# Optical blood flow monitoring in humans with SNSPDs and high-density SPADs

**DOI:** 10.1101/2025.06.08.25329202

**Authors:** Carsi Kim, Christopher H. Moore, Chien-Sing Poon, Michael A. Wayne, Paul Mos, Arin Ulku, Timothy M. Rambo, Aaron J. Miller, Claudio Bruschini, Edoardo Charbon, Ulas Sunar

## Abstract

Continuous, noninvasive monitoring of cerebral blood flow (CBF) is vital for neurocritical care. Diffuse correlation spectroscopy (DCS) enables assessment of microvascular blood flow by analyzing speckle intensity fluctuations of near-infrared light. In this review, we summarize recent advances in TD-DCS using superconducting nanowire single-photon detectors (SNSPDs) at 1064 nm, as well as complementary developments in high-density CW-DCS systems using single-photon avalanche diode (SPAD) cameras. Time-gated photon detection improves depth sensitivity in TD-DCS, and the use of longer wavelengths provides advantages in tissue penetration, photon throughput, and safety margin under ANSI exposure limits. Clinically feasible SPAD-based implementations, while lacking time-of-flight resolution, enable large signal-to-noise ratio gains via massive pixel averaging and offer a room-temperature, scalable path to high-density optical tissue monitoring. Together, these developments highlight a growing set of technologies for clinical applications, including bedside brain monitoring in neurocritical care. We conclude with practical guidance on detector technologies, gating strategies, system packaging, and briefly discuss interferometric DCS and speckle contrast optical spectroscopy (SCOS) as synergistic extensions for high-resolution and high-coverage imaging.

## 1. Introduction

Diffuse correlation spectroscopy (DCS) is an optical technique for noninvasive assessment of microvascular blood flow. It measures speckle intensity fluctuations from coherent near-infrared light propagating through scattering tissue, enabling continuous monitoring of tissue blood flow *in vivo* [1–4]. For a comprehensive review of diffuse optical methods for blood flow and tissue monitoring, the reader is referred to Durduran et al. [4], and for a current status of DCS the reader is directed to Carp et al [5]. Conventional continuous-wave DCS (CW-DCS) faces limitations in depth sensitivity, signal contamination from extracerebral tissues, and reduced signal-to-noise ratio (SNR) at longer source-detector (SD) separations [6–10].

Sutin et al. [11] from the Boas group, introduced a novel time-domain (pathlength-resolved) TD-DCS method. TD-DCS addresses these limitations by using pulsed lasers and time-resolved photon detection. By selecting photons based on their time-of-flight (TOF), TD-DCS provides pathlength-resolved sensitivity that improves depth discrimination and cerebral specificity. Operation at shortwave infrared (SWIR) wavelengths, such as 1064 nm, further enhances penetration and photon throughput because of reduced scattering and higher maximum permissible exposure under ANSI safety guidelines [5]. Together with superconducting nanowire single-photon detectors (SNSPDs) and time-correlated single-photon counting (TCSPC) electronics, TD-DCS can be implemented in compact, fiber-coupled probes suitable for bedside neuromonitoring. Feasibility has been demonstrated under physiologic manipulations (e.g., pressure modulation), in neurocritical care settings, and during physiological maneuvers such as Valsalva or a breath-hold maneuver [5,12–16]. For a detailed review of TD-DCS, covering theoretical foundations through initial human studies, the reader is referred to Tamborini et al. [17].

In parallel, single-photon avalanche diode (SPAD) arrays have emerged as a complementary approach to improve SNR in CW-DCS. Although current SPAD systems do not provide TOF resolution, their massively parallel pixels enable efficient multi-speckle averaging, yielding large SNR gains without high laser powers. SPAD-based systems operate at room temperature and support flexible, modular, and clinically adaptable neuro-monitoring platforms [18– 21]. They have already been applied in several blood-flow contexts [19,21–30]. While present-generation SPAD devices offer only coarse autocorrelation times and large gates, ongoing advances in temporal resolution and 1064 nm sensitivity could expand their capabilities.

This review summarizes current TD-DCS implementations using SNSPDs at 1064 nm (and ∼785 nm where relevant) and SPAD-based CW-DCS systems, comparing their strengths, trade-offs, and translational considerations. We conclude with recommendations on detector selection, gating methods, and system integration, and note (iDCS) and speckle contrast optical spectroscopy (SCOS) as complementary approaches for broad-coverage, depth-resolved clinical monitoring.

## 2. Methods

### 2.1 Model for Tissue Blood Flow Index

When coherent light enters tissue, it undergoes multiple scattering events, where moving red blood cells result in temporal fluctuations in the detected speckle intensity. The temporal autocorrelation of these intensity fluctuations reflects the underlying blood flow dynamics [1–3,5,8,11,31–36]. The detected photon intensity autocorrelation function, *g*_2_(*t*_*s*_, *τ*), is defined as, 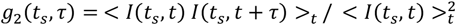, where *t*_*s*_ is the TOF, or “micro-time”, which is proportional to the photon pathlength through the medium, *τ* is the correlation delay (lag time), <. >_*t*_ denotes averaging over macro-time, *t* is the absolute photon arrival time from the start of the measurement. In TD or time-gated DCS, this time-resolved autocorrelation function is typically modeled using the Siegert relation, *g*_2_(*t*_*s*_, *τ*) = 1 + *β*| *g*_1_(*t*_*s*_, *τ*)| ^2^, where *g*_1_ is the normalized electric field autocorrelation function, which encodes information about scatterer motion, and *β* is the coherence factor. In the diffusion approximation, *g*_1_(*s, τ*) is expressed as an integral over photon path lengths *s*, where *s* is the total optical path length traveled by the photon through the scattering medium, corresponding to its TOF and related to the tissue geometry and scattering properties, and the model becomes 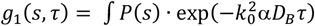 *ds*, where *P*(*s*) is the photon path length distribution from the temporal point spread function (TPSF), *D*_*B*_ is the effective Brownian diffusion coefficient, and *k*_0_ = 2πn/λ is the optical wavenumber with *n* being the index of refraction, and *λ* the wavelength. The blood flow index (BFI) is then estimated as α*D*_*B*_ where α is the fraction of dynamic scatterers. Unlike CW-DCS, TD-DCS models *g*_2_ as a function of pathlength-resolved photon dynamics, offering enhanced depth sensitivity [11,15,32,37– 41].

### 2.2 Instrumentation

Fig.1(A) depicts a simplified TD-DCS setup. A long coherence laser emits pulses of light at a specified repetition rate (e.g., 80 MHz). The laser’s sync signal is coupled to a TCSPC module where the detected photons are tagged to each laser pulse. The laser source is typically transmitted into the tissue through a multimode fiber (MMF). Photons exiting from the tissue are detected by a single mode fiber coupled to a single photon detector. Currently the most common detectors used in TD-DCS are SNSPDs or SPADs [14,39]. Next, a TCSPC module registers detected photons to timed laser pulses, generating a TPSF curve. From this TPSF curve different photon arrival time gates can be chosen based on the depth or pathlength of interest. Photons arriving before the peak (assuming infinitesimal instrument response function (IRF)) are referred to as early gate (EG) photons and signify travel through the superficial layers of tissue. Photons arriving near the tail end of the TPSF peak are quantified as late gate (LG) photons and indicate deeper penetration due to longer photon arrival time. These gated photons are then autocorrelated as briefly described in the model section to generate *g*_2_, which is then used to calculate BFI. A general SPAD-based DCS system is shown in Fig.1(B). In this setup, a coherent laser is delivered to the tissue via a MMF. The scattered light is collected by a MMF and directed onto a SPAD array detector. The fiber is set at a specific distance away from the SPAD sensor to match speckle size to the pixel active area. The SPAD array captures multiple spatially independent speckle patterns in parallel allowing for higher SNR [19,30] . Each pixel detects individual photon events over time, enabling the tracking of dynamic speckles that change as blood cells move within the tissue. The time-varying speckle patterns are then processed on a field programmable gate array (FPGA) or through software to eventually calculate BFI [18,19,21]

**Fig. 1.**
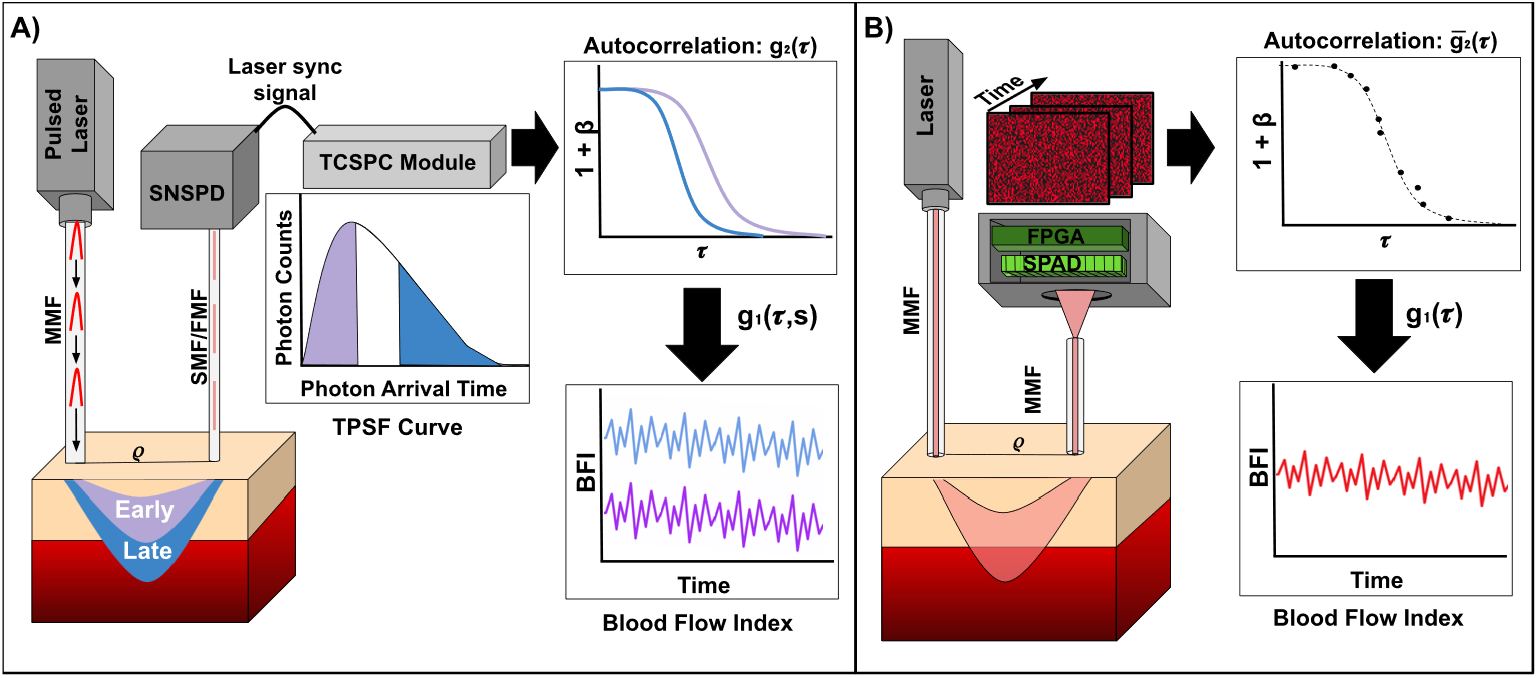
The schematic illustrates (A) TD-DCS and (B) SPAD-based multi-pixel DCS (mDCS) setups. (A) A pulsed laser delivers coherent light into tissue. The time-correlated single-photon counting (TCSPC) module records photon time-of-flight to generate a temporal point spread function (TPSF). Time-gating selects photons by arrival time-early (purple) and late (blue) gates. From these gated photons, the autocorrelation function is computed as a function of photon path lengths, s, to estimate the blood flow index (BFI). (B) The mDCS workflow using a SPAD array in continuous wave. “Each pixel” behaves as detector. Then the electric field autocorrelation is derived via the Siegert relation. The resulting g2(*τ*) is then fitted to a diffusion model to estimate the blood flow index (BFI).

### 2.3 Detector Specifications: SNSPDs & SPAD Camera

For TD-DCS, we employed two representative system configurations, each based on a 4-channel TCSPC unit (HydraHarp400, PicoQuant, Berlin, Germany). In one setup, detection was performed with four SNSPD channels coupled to a 1064 nm pulsed seed laser (QDLaser, Japan) and amplifier (Cybel, USA). In the other, a 1064 nm pulsed laser (PicoQuant, Germany) was interfaced directly with multimode detection fibers. In both cases, the probe design included a prism-mounted diffuser to expand the illumination beam (∼4.5 mm), and power levels were maintained below ANSI safety limits, with photon count rates of ∼10^7^ cps per channel. These configurations illustrate the typical experimental approaches for TD-DCS.

The SPAD camera array used for this study was a SwissSPAD3, which is a 500 x 500 binary, dual-gate CMOS SPAD array split into two 250 x 500 halves. Its readout is virtually noiseless, thanks to the direct in-pixel photon-to-digital conversion. This allows operation at very high frame rates with no penalty. Each half requires its own FPGA daughter board and only one half was used here. The camera has a pixel pitch of 16.38 µm with a SPAD active area of 6 μm diameter per pixel [22]. Each SPAD has a breakdown voltage of ∼23 V and was operated at 28 V for an excess bias voltage of ∼5 V. At an operating wavelength of 785 nm this allows for a photon detection probability of ∼13%. With the camera’s maximum framerate of close to 100 Kfps, the FPGA can calculate the autocorrelation with a minimum lag time of nearly 10^-5^ s [19].

## 3. Validation of TD-DCS with SNSPDs at 1064nm

It has been shown that by selecting the SD separation and time-gate (pathlength-resolve), one can achieve higher contrast to changes in deeper layer dynamics, such as cerebral blood flow (CBF) change in the cortex induced by functional activation [11,12,32,42]. TD-DCS uses a pulsed laser, which may have a limited laser coherence length (*L*_*C*_), thus the measurement is affected by the instrument response function (IRF) and *L*_*C*_. To obtain highly sensitive pathlength-selected DCS measurement of deep CBF, several experimental factors including IRF, *L*_*C*_, gate opening time, gate width, and SD separation should be considered in the experimental design and data analysis. Cheng *et al*. has developed an analytical model of time-resolved *g*_2_ containing IRF and *L*_*C*_ factors and applied it to a semi-infinite medium [43]. Furthermore, the IRF effects were investigated [41] and a model was proposed to extract the blood flow from the gated intensity autocorrelation function [44].

Here, we show representative data to demonstrate *in-vivo* SNR advantage at 1064 nm over traditional, lab-standard 785 nm CW-DCS, head-of-bed (HOB) protocols in human subjects, pressure modulation studies, and clinical neuro-ICU patient monitoring are shown by Poon et al. Along the lines of previous work on CW-SPAD using human subjects, such as mental tasks [45], here we show the feasibility for monitoring blood flow through a hand grip and HOB protocols performed in parallel with a CW-DCS (lab-standard) system.

### 3.1 Increased Signal-to-Noise Ratio at 1064 nm

The Mie scattering model predicts that the scattering parameter decreases as a power law with respect to wavelength [48]. Since optical attenuation is strongly dependent on optical scattering, this spectral behavior leads to significant reduction in light attenuation and deeper light penetration at longer wavelengths [49]. Fig.2(A) shows the transmittance spectra for freshly harvested rat brain, cranial bone, and skin tissue [50]. This data confirms that transmission increases with wavelength due to the reduction in scattering. However, while transmittance continues to rise beyond 1064 nm, water absorption also increases, limiting practical gains in SNR for human cerebral applications beyond this wavelength. As such, 1064 nm represents an optimal spectral region for deep cerebral imaging, balancing reduced scattering and manageable absorption. Accordingly, systems developed by multiple groups have focused on this wavelength for optimal depth sensitivity [16,33,37,51]. To validate this spectral advantage in practice, DCS measurements were performed at 780 nm and 1064 nm using SNSPDs with high efficiency at 1064 nm. As shown in Fig.2(B), measurements demonstrated an approximately 9-fold increase in SNR at 1064 nm compared to 780 nm at a 2.8 cm SD separation in the brain. This improvement in photon counts is especially valuable in TD-DCS, where late-arriving photons are sparse and critical for probing deeper tissues.

**Fig. 2.**
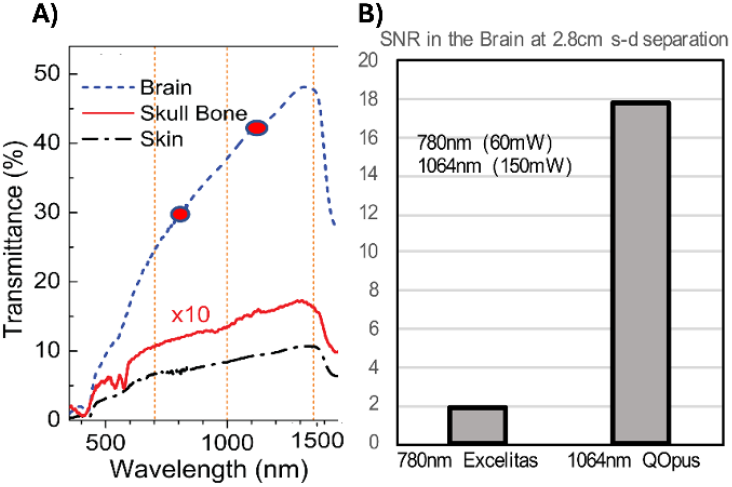
(A) Increased transmission in the rat brain with increasing wavelength. Reproduced with permission from Tymish Y. Ohulchanskyy, Junle Qu, Liwei Liu, et al, Journal of Biophotonics, © 2018 Wiley-VCH GmbH. (B) Measured SNR in comparison with 780nm Excelitas and 1064nm Quantum Opus SNSPDs at 2.8 cm source-detector (s-d) separation on a human head.

### 3.2 Human Head-of-Bed Tilt Protocol

We first worked in CW-DCS mode at 1064 nm with the established, clinically relevant protocol of HOB manipulation to check the practical signal gain and demonstrate the feasibility of clinical translation with improved SNR in the human brain. HOB manipulation has been used to evaluate cerebrovascular health in TBI and stroke patients, and CW-DCS at 785 nm using Excelitas detectors has been used successfully for this protocol in these patients and healthy subjects [8,52–56]. Twelve healthy subjects were recruited, and measurements were taken from the prefrontal cortex (between Fp1 and F7 of the EEG 10/20 system) while a motorized bed was tilted to 30 degrees and then returned to 0 degrees to modulate CBF. The 4.5 μm single mode fiber was placed at 2.8 cm away from the source and is connected to SNSPDs. Data was sampled at ∼0.3 Hz. Representative *g*_2_ curves show clear differences between baseline and tilted conditions, seen in Fig.3(A). The average BFI increased during the tilt, p< 0.001, rank sum test, seen in Fig.3(B), with group level box plots showing overall responses across subjects (Fig.3C). On average, the tilt produced a ∼24.2±16.2% increase in BFI (Fig.3(D)). These results align well with the previous HOB studies performed by CW-DCS at 785nm, where an increase in BFI is seen when subjects are supine compared to upright [8,35], and that SNSPDs can reliably obtain CBF changes at 1064nm in humans in a clinically relevant protocol, with higher SNR and slightly longer SD separation (2.5 cm vs 2.8 cm).

**Fig. 3.**
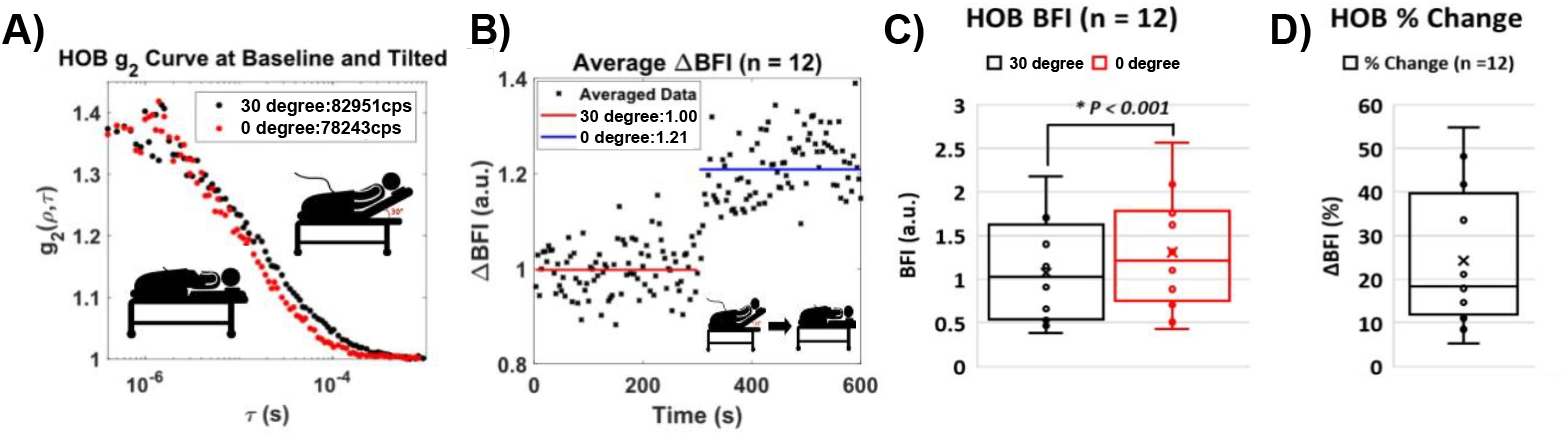
The head of bed (HOB) experiment is performed on 12 subjects. (A) A representative g2 curve between baseline, black, and tilted, red. (B) Data compared between the group average baseline and tilted cases and a difference was observed in the blood flow index (BFI) with a p<0.001 using rank sum test. (C) Comparison of average BFI during baseline and tilted. (D) Percent change of BFI when transitioning between baseline and tilted.

### 3.3 Pressure Modulation Experiment

To validate the TD-DCS system with SNSPDs *in vivo* a typical pressure modulation was performed on the head, and then a staged pressure modulation where pressure differences were set into timed increments, similar to previous work [13,33,37,38,51,57]. A typical protocol for TD-DCS has been pressure modulation, where the probe is pressed on the forehead to reduce blood flow seen from the superficial layer, predominantly the scalp. A pressure modulation protocol on the scalp to occlude the scalp blood flow was done by controlling the pressure application, around 50mmHg, by pressing the optical probe for 60s after a 60s baseline, as seen in Fig.4(A). CW and EG had higher changes (63%, 76%, respectively) vs 30% change in LG, as expected since LG photons have less sensitivity to scalp (Fig. 4(B)). Overall, these results show LG photons have high sensitivity to deeper flow.

**Fig. 4.**
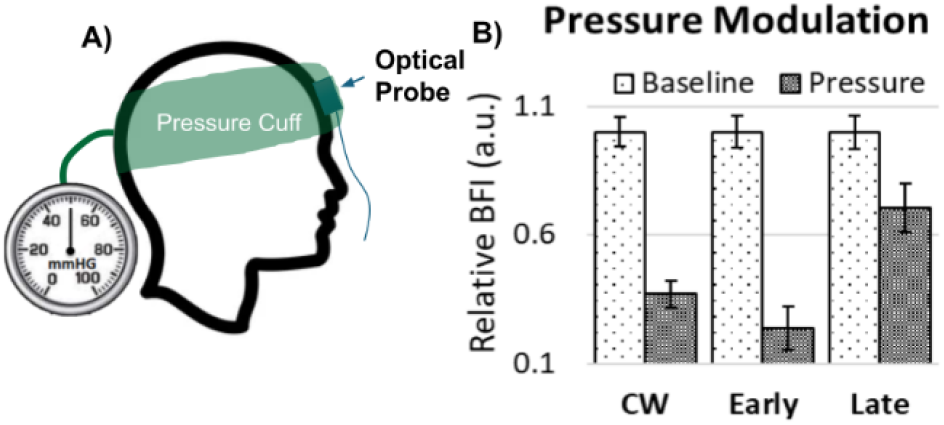
(A) Pressure modulation on scalp. Scalp is pressed hard (∼50 mmHg) to reduce the blood flow in the scalp. (B) Late-gate (LG) photons show less change compared to continuous-wave (CW) and early-gate (EG) groups.

### 3.4 Clinical NeuroICU Case Study

Beyond validation protocols, the development of SNSPD technology itself is rapidly advancing, bringing TD-DCS closer to clinical translation. Several studies have already validated TD-DCS in-vivo with SNSPDs in healthy subjects, showing potential for clinical translation [15,16,39]. The first clinical application of SNSPD-based TD-DCS was performed by Poon et al [51] using an 8 channel TD system shown in Fig.5(A). This setup is large, however, more compact commercial SNSPD systems can support up to 32 detection channels and are smaller in size, Fig.5(B). In this clinical study, TD-DCS at 1064 nm was used in a case study for a patient with severe traumatic brain injury in a neuro-ICU setting. Gating was used in the TD-DCS measurements to separate superficial with deeper layer blood flow. Multiple parameters were characterized in this study, including IRF, TPSF, EG, LG, g2 curves, and CBF. EG was taken as -100pf from the peak with a width of 10ps, and LG was taken as +650 ps from the peak with a width of 800 ps. IRF was taken by pointing the source and detector fibers to each other with a thin piece of Teflon in between. EG and LG BFI values were compared to invasive thermal diffusion flowmetry (TDF) seen in black and red, respectively in Fig.7(A,B,C). The BFI and TDF values yielded strong correlations, ρ = 0.67 for EG and ρ = 0.76 for LG. The TD-DCS system also operated at a high sampling rate of 50 Hz, allowing for the detection of pulsatile blood flow, as seen in Fig.7(D). Of particular interest was the TD-DCS system’s ability to capture clear responses to clinical events such as sedation, withdrawal, neurological examination, and sedation resumption. In Fig.7(A,B,C) the three green markers indicate key clinical events: (1) sedation stopped, (2) neurological exam performed, and (3) sedation resumed. These differing responses between the markers are seen in both TD-DCS and in the TDF perfusion measurements. These results highlight the clinical viability of SNSPD-based TD-DCS, demonstrating its ability to reliably track CBF in real time with high sensitivity and agreement to the invasive gold-standard technique.

**Fig. 5.**
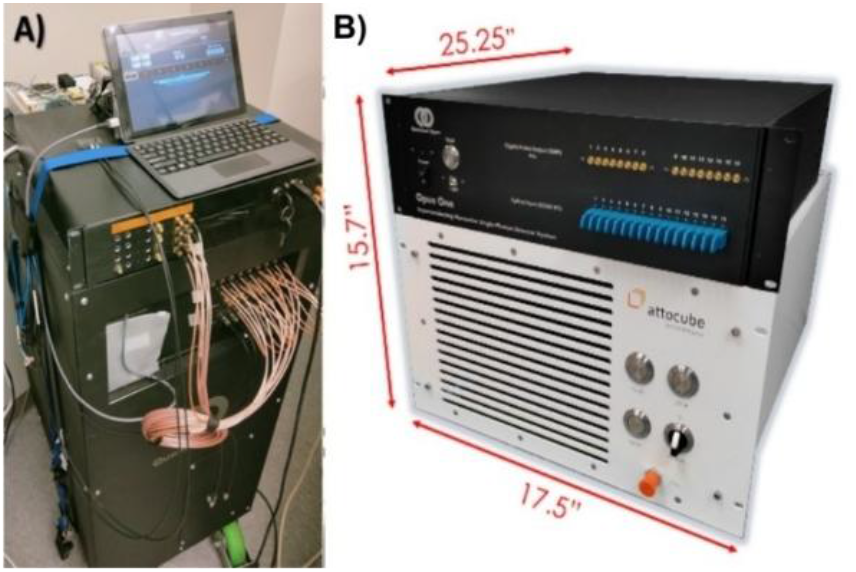
(A) Clinically translated version, 8-channel. (B) A complete 32-channel SNSPD system in a 7U rack mountable package using only commercial off-the-shelf components

**Fig. 6.**
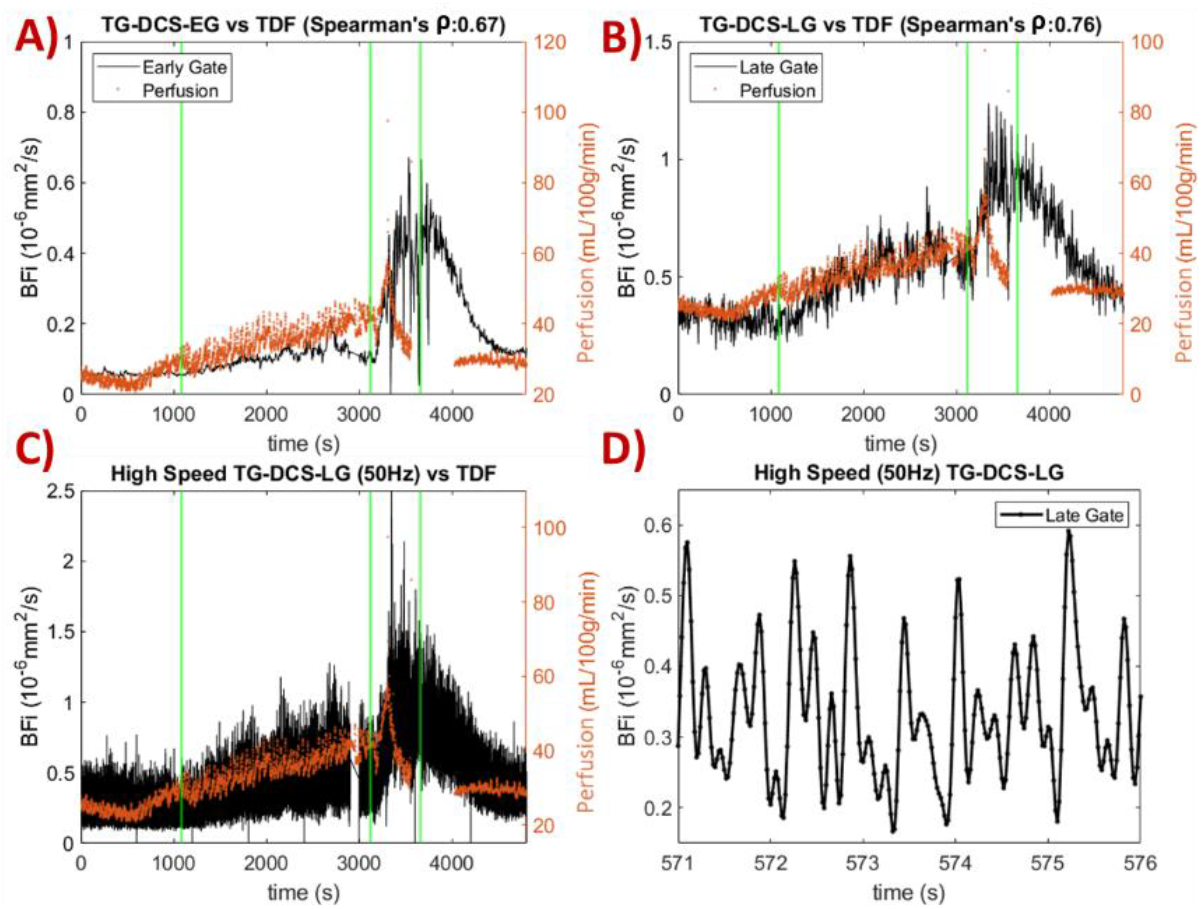
(A) Early-gate blood flow index (EG-BFI). (B) Late-gate BFI (LG-BFI) compared to invasive thermal diffusion flowmetry (TDF) cerebral blood flow (CBF), with Spearman’s correlation coefficients of 0.67 and 0.76, respectively. (C) The 50 Hz LG-BFI signal captures distinct fluctuations associated with pulsatile blood flow. (D) A zoomed-in view of the 50 Hz BFI trace highlights pulsatility. The three green markers indicate key clinical events: (1) sedation stopped, (2) neurological exam performed, and (3) sedation resumed. Adapted from (Poon et al, 2022) [51].

**Fig. 7.**
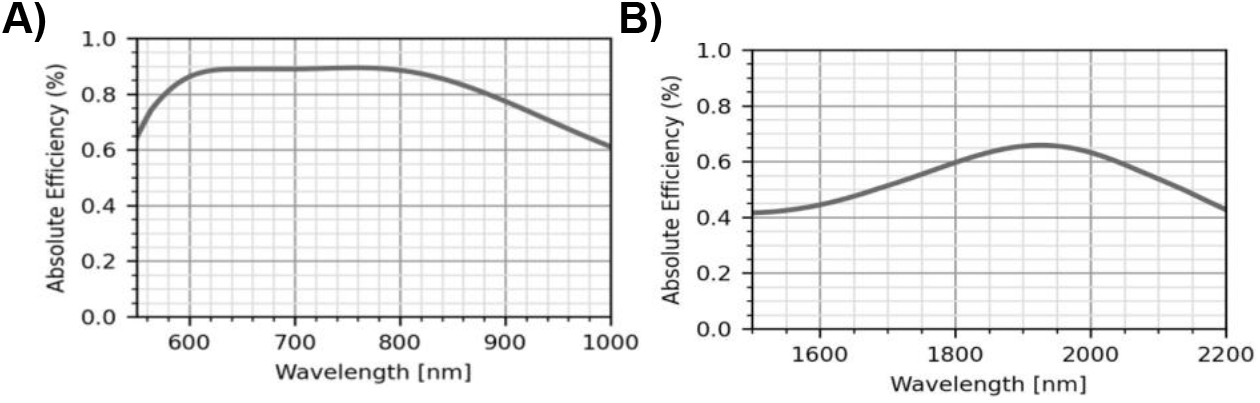
Typical detector performance curves reaching into the (A) visible, near infrared, and (B) infrared regimes based on Quantum Opus characterizations.

While SNSPDs offer higher performance for many scientific applications, one key barrier to widespread adoption is that they require operation at cryogenic temperatures below 4 K. Pre-existing cryogenic technology can be packaged in a way that is roughly compatible with clinical settings, but the result is still bulky, noisy, and requires a large amount of power. New developments in commercial off-the-shelf cryogenic components can facilitate repackaging of the detectors into a smaller and lower power system, such as a 7U, all-in, fully-rack mountable SNSPD system with integrated vacuum pump and electronics [58]. By incorporating such a high number of channels into a significantly lower size, weight and power system, SNSPD TD-DCS would be able to support many measurement channels without imposing on the clinical environment. Another key advantage of superconducting detectors is their broadband optical response, spanning from UV [59] to mid-infrared [60] (Fig.7). While commercial SNSPDs were originally designed for near infrared and telecom wavelengths, they are now expanding through the visible spectrum and 2000 nm regimes. High performance detection at 2000 nm can allow for measurements with less scattering albeit higher absorption.

Furthermore, SNSPDs have seen improvements in their temporal characteristics with higher sampling rates and precision [61]. Following a detection event, recovery of the bias current in the nanowire structure dictates the detector’s maximum count rate, which could exceed 100 MHz, an order of magnitude higher than conventional SPADs and earlier SNSPDs. This allows for faster data acquisition, or operation at lower laser peak power with higher repetition rate lasers. This can potentially eliminate the need for optical amplifiers. Additionally, with respect to parallel detection, current SNSPD implementations have been limited in terms of array size and pixel density. Recent breakthroughs, such as the development of a 400,000-pixel SNSPD camera by the NIST group, demonstrate a major step forward, offering a 400× improvement over previous kilo pixel limits [62]. As SNSPDs continue to advance in form factor, bandwidth, timing precision and scalability to high-density pixels, they are becoming increasingly practical for clinical translation.

## 4. Validation of High-Density SPAD-Based mDCS Measurements

To explore scalable alternatives to SNSPD-based TD-DCS, we implemented a DCS system using a high-density SPAD array operating in CW mode, as similar in architecture to other recently reported SPAD-DCS systems [19,21,23,30,63–65]. Our SPAD camera, SwissSPAD3 (SS3), fabricated in CMOS technology, is shown in Fig. 8. The setup includes a 3D-printed fiber holder, a SS3 daughterboard containing the SPAD array PCB, a motherboard with intermediate components, and a dedicated FPGA daughterboard responsible for high-speed acquisition. This system does not use active gating but instead performs post-processing on photon arrival time windows to compute intensity-based autocorrelations. By leveraging an FPGA-based acquisition pipeline, we achieved a ∼500-fold improvement in SNR compared to a conventional single-detector DCS system [19]. This high level of parallelization enables sensitive flow detection in deeper tissue and within shorter acquisition windows and facilitates monitoring of rapid hemodynamic changes in deep tissue. To validate this system in vivo, we conducted human subject measurements during both handgrip and head-of-bed (HOB) protocols. These measurements reproduced expected physiological blood flow trends, consistent with prior CW-DCS findings.

**Fig. 8:**
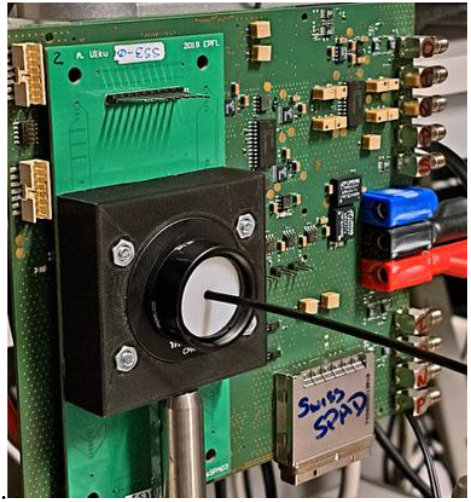
Image of the SwissSPAD3 (SS3) setup with fiber-coupled detection. The system includes a SPAD array on the daughterboard (main PCB), a motherboard with intermediate routing and support components, and an FPGA daughterboard that handles real-time acquisition and processing.

### 4.1 Hand-Grip Protocol

To assess SPAD system performance *in vivo*, we performed simultaneous CW-DCS and SPAD-based mDCS measurements during a handgrip exercise in a healthy subject. The protocol consisted of a 100 s baseline, 60 s isometric handgrip, and 100 s recovery. The primary physiological response to isometric exercise is an increase in local blood flow due to vasodilation, driven by increased metabolic demand and accumulation of lactate and CO_2_. Sympathetic vasoconstriction is overridden locally, leading to active hyperemia. Upon release, a transient overshoot in blood flow, as post-exercise hyperemia, occurs due to residual vasodilation and the removal of mechanical compression. Figure 9A shows the BFI time course measured by both systems. The SPAD-based mDCS closely matched the CW-DCS signal, demonstrating a clear increase during contraction and a decline during recovery. This alignment confirms the SPAD array’s ability to replicate expected physiological trends.

**Fig. 9.**
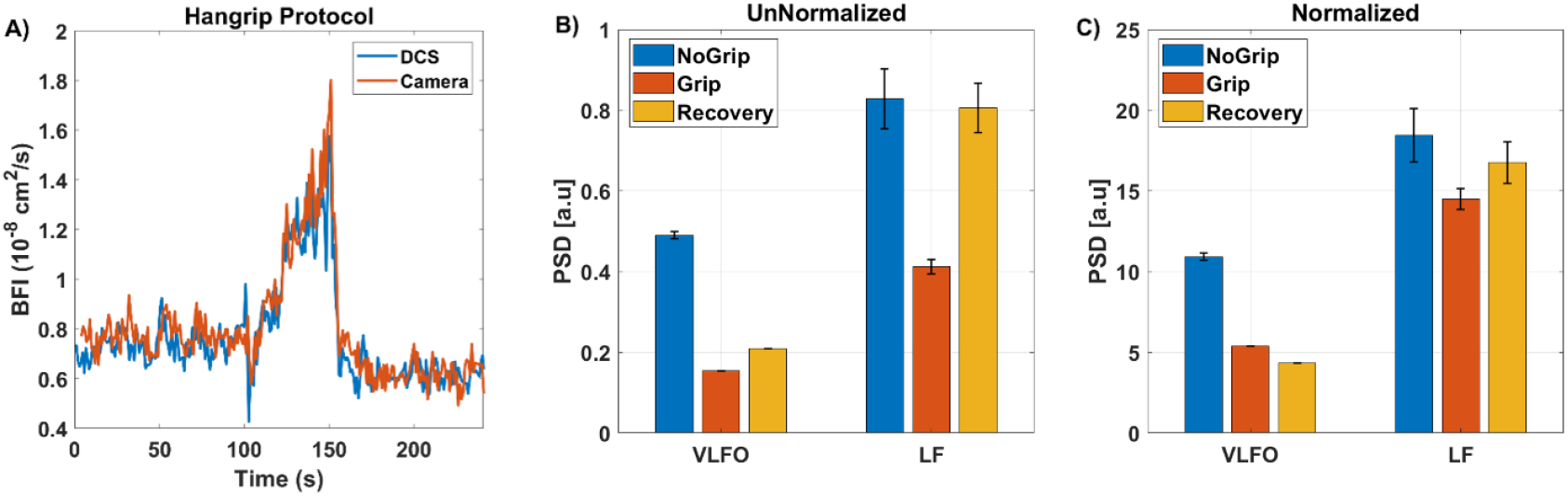
Blood flow measurements during a handgrip protocol using SPAD-based mDCS. The protocol included 100 s of rest, 60 s of handgrip, and 100 s of recovery. The probe was placed on the inner forearm and secured with Velcro or adhesive wrap. (A) Time-series plot of blood flow index (BFI), showing increased perfusion during grip followed by a post-exercise drop. CW-DCS is shown in blue, and SPAD-based mDCS in red. (B) Power spectral density (PSD) in very low frequency (VLF: 0.02–0.07 Hz) and low frequency (LF: 0.07–0.20 Hz) bands, computed from unnormalized BFI. (C) PSD results normalized by total power (area under the PSD curve) to highlight relative changes in spectral content across conditions. Error bars represent standard deviation across frequency bins within each band.

To explore slow hemodynamic fluctuations, we analyzed low-frequency oscillations (LFOs) in the BFI signal. Prior work in skeletal muscle and skin has linked LFOs to distinct physiological mechanisms. The LF band (0.07-0.20 Hz) reflects myogenic regulation of vascular tone during contraction while the VLF band (0.02-0.07 Hz) is associated with neurogenic and endothelial modulation during recovery [66,67]. Figures 9B and 9C show the PSD values for VLF and LF bands in absolute and normalized forms, respectively. The error bars reflect variability across frequency bins. In the unnormalized PSD (Fig. 9B), the grip period shows increased BFI and decrease PSD power in both LFs and VLF, most likely due to the shift in baseline regulation to metabolic control, consistent with prior blood flow studies during a stimulus [61,68,69]. In the normalized PSD (Fig. 9C), relative power distributions shift, emphasizing spectral shape changes rather than total power. Together, these analyses confirm the physiological fidelity of SPAD-based mDCS during exercise-induced vascular dynamics.

### 4.2 Human Head-of-Bed Tilt Protocol

A HOB protocol, similar to the DCS SNSPD system, was performed for the SPAD system but for different time durations. This experiment consisted of a healthy subject’s data for a HOB protocol of 2 minutes supine and 2 minutes tilted upright for 30 degrees.

To further evaluate the SPAD-based mDCS system for cerebral applications, we conducted a head-of-bed (HOB) tilt protocol in a healthy subject. The subject was monitored at a 2.5 cm SD separation using an optical probe placed on the forehead. The protocol consisted of 150 seconds in the upright position (30°-tilt), followed by 150 seconds in the supine position. Figure 10A shows the BFI time series throughout the protocol. As expected, blood flow increased when the subject was tilted supine, a pattern generally associated with changes in posture. Figure 10B presents the average BFI values for both positions, revealing a ∼22% increase in the supine condition. This trend aligns with prior measurements using CW-DCS at 1064 nm (Fig. 5) and with published results using traditional DCS systems at 785 nm [8,35]. Although cerebral autoregulation typically maintains stable blood flow during mild postural changes, we observed a consistent ∼15-20% change in BFI between upright and supine conditions. This likely reflects contamination from superficial scalp blood flow, which increases in the supine position due to venous pooling. Such extracerebral contributions are well known to affect CW-DCS signals in the absence of time-gating (pathlength selection) or multi-layer modeling, as observed in prior work using traditional DCS systems [8,35]. These results emphasize the importance of developing time-resolved SPAD-based systems and/or applying data correction strategies to improve cerebral specificity in future work.

**Fig. 10.**
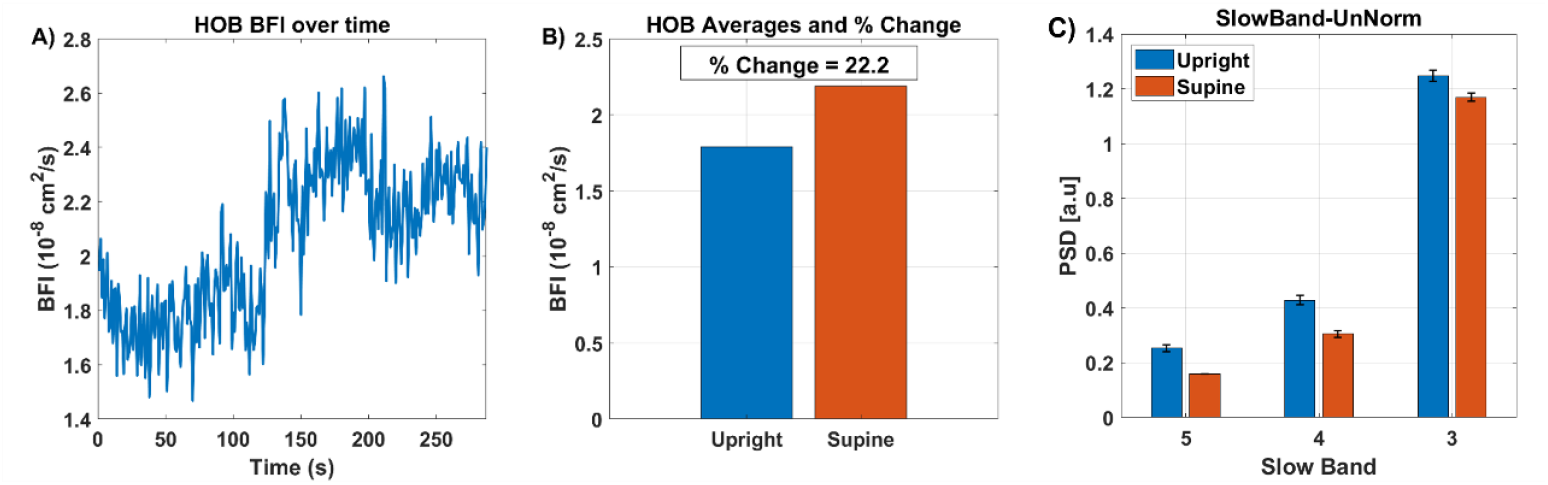
Blood flow measurement during a head-of-bed (HOB) tilt protocol using SPAD-based mDCS at a 2.5 cm source–detector separation. The protocol included 150 s upright (30° tilt) followed by 150 s supine. (A) Time-series of blood flow index (BFI), showing increased cerebral perfusion in the upright position. (B) Averaged BFI values for upright and supine conditions, with a calculated ∼22% increase when supine. (C) Power spectral density (PSD) in three low-frequency bands: slow-5 (0.01-0.027 Hz), slow-4 (0.027-0.073 Hz), and slow-3 (0.073-0.198 Hz), for upright and supine positions. All PSD values are normalized by total power, and error bars reflect standard deviation across frequency bins.

To assess slow hemodynamic oscillations, we computed power spectral density (PSD) across three established low-frequency bands: slow-5 (0.01-0.027 Hz), slow-4 (0.027-0.073 Hz), and slow-3 (0.073-0.198 Hz) [70–78]. Figure 10C displays unnormalized PSD for the upright and supine positions. While variability exists due to the short acquisition window, the observed spectral trends are consistent with physiologic modulation of cerebral and extracerebral blood flow dynamics, where healthy patients are theorized to have little differences between supine and upright due to proper cerebral autoregulation [8,79–81]. These shifts may reflect systemic contributions or superficial vascular tone and are expected to be more pronounced in patients with impaired autoregulation, such as those with acute brain injury.

Overall, these results support the feasibility of SPAD-based CW-DCS for cerebral blood flow monitoring and highlight a clear path toward the future systems that combine room-temperature scalability with depth-sensitive detection through time-gating or model-based correction.

## 5. Discussion and Outlook

This review summarizes recent developments for blood flow monitoring using SNSPDs and high-density SPAD array technologies. These advances reinforce the value of time-gated photon detection for enhancing depth sensitivity for human translation, particularly at 1064 nm where reduced scattering and higher permissible power improve signal quality relative to traditional 785 nm CW-DCS systems. MC simulations and human data support that LG photons are more sensitive to deeper cerebral layers than EG or CW approaches, addressing a major limitation of CW-DCS, which lacks pathlength selectivity. SNSPD-based TD-DCS systems enable high temporal resolution and strong SNR due to their exceptional detection characteristics. Physiological manipulations such as a HOB tilt and pressure modulation have shown consistent BFI changes that align with expected autoregulatory responses and prior literature. Additionally, the observed correlation between LG BFI and invasive thermal diffusion flowmetry supports the translational relevance of TD-DCS.

SPAD-based systems, while currently operating in CW mode, represent a scalable and portable solution for high-density neuro-monitoring. Although they do not yet offer true TOF resolution, recent advances in multi-channel InGaAs/InP SPAD technology, such as those by the Charbon group [82], have demonstrated ∼35% photon detection efficiency at 1064 nm, ∼100 ps jitter, and dark count rates on the order of 10^3^ cps, supporting the feasibility of TD-DCS in the shortwave infrared. Parallel efforts by MIT Lincoln Lab have produced 3D-stacked SPAD imagers with high fill factor and integrated gating, designed for applications at 1064 nm [83]. Together, these advances point toward the potential for gated or hybrid SPAD-based systems that combine the scalability with partial TOF sensitivity, bridging the gap toward time-domain performance in a compact, room-temperature platforms.

Another emerging approach is the development of interferometric diffuse correlation spectroscopy (iDCS), which aims to achieve TOF capability without reliance on pulsed lasers or cryogenic detectors, as initially demonstrated by Borycki et al [84]. Recent iDCS systems have demonstrated enhanced sensitivity to deep tissue through pathlength-selective coherent gain, enabling depth-resolved blood flow measurement with simplified instrumentation [85]. For example, Zhao *et al*. demonstrated a scalable iDCS system with over 200 channels and electronically tunable coherence gating, which reduced scalp contamination and enabled TOF-resolved imaging [86]. Additional interferometric variants have been reported, including functional interferometric diffuse wave spectroscopy (iDWS) using tunable coherence gating [25], coherent amplification using CMOS sensor arrays [19,21], and a scalable interferometric system that leverages high-speed, 2D CMOS cameras for depth-sensitive measurement in a continuous-wave framework [27]. Very recently, iDCS has advanced toward clinical translation in a neurocritical care setting [87]. Collectively, these coherence-based techniques offer scalable, room-temperature alternatives to TD-DCS, potentially mitigating the cost, size, and cooling requirements of cryogenic systems while preserving key benefits such as depth selectivity and TOF-like sensitivity.

Despite its advantages, TD-DCS faces several technical and translational limitations. These challenges include the difficulty of balancing short laser pulse width and long coherence length, managing signal attenuation at deeper gates, and optimizing fiber geometry and coherence factor. SNSPDs offer unmatched performance metrics including near-unity quantum efficiency, <3 ps timing jitter, ultralow dark counts, and broadband spectral response but require cryogenic operation and sophisticated packaging, limiting portability and clinical translation. Moreover, system-level calibration of the IRF and standardization across platforms remain active areas of development. SNSPD-based TD-DCS remains attractive for ultra-sensitive applications, particularly at 1064 nm where its performance in photon-starved clinical settings are unmatched. As large-format SNSPD arrays become viable alternatives or complements to SPAD arrays in TD-DCS, their role may expand from single-point probes to high-density imaging platforms, particularly for deep-tissue or functional neuroimaging where absolute sensitivity is critical. Further work is needed to evaluate the trade-offs in cryogenic complexity, integration, and cost, but SNSPD-based time-domain diffuse optics could provide excellent performance in photon-starved conditions and unlock deeper imaging capabilities in neuroscience and neuroimaging applications.

An additional recent approach for noninvasive blood flow monitoring is diffuse speckle contrast analysis (DSCA) [88–90], also referred to as speckle contrast optical spectroscopy (SCOS) [88,91–94]. DSCA/SCOS uses high-resolution CMOS cameras to track spatiotemporal speckle fluctuations. Although the DSCA/SCOS approach lacks the ability to recover the full *g*_2_(*τ*) curve, it benefits from low cost, wide-field, high-density coverage, and ease of instrumentation [88,91–94]. SCOS systems can be scaled to multichannel arrays and have demonstrated sensitivity to cortical hemodynamics with high spatial and temporal resolution [95,96]. Ongoing improvements in fiber coupling, speckle processing, and noise correction are expanding the range of SCOS applications. Meanwhile, SPAD-based DCS provides full *g*_2_(*τ*) recovery and is expected to be more sensitive to cerebral blood flow index estimation. Assuming optical parameters are known or estimated a priori, this enables quantitative monitoring, which may be required in several applications such as stroke and hydrocephalus where monitoring for absolute ischemia thresholds are clinically relevant for guiding interventions. Recent improvements in SPAD sensor design, such as 3D-stacked CMOS SPAD arrays from the Henderson group, have achieved ∼1 µs minimum lag time, 100% fill factor, and photon detection efficiencies exceeding 50%, all within compact, room-temperature packages [23]. These advances are expected to accelerate SPAD system translation for human neuro-monitoring.

Looking ahead, the next generation of optical blood flow monitoring systems will likely involve hybrid architectures that combine SNSPDs, SPADs, and CMOS technologies with each contributing distinct advantages. Multi-channel SNSPD arrays may enable depth-resolved imaging with exceptional sensitivity, particularly for absolute TD-DCS and functional neuroimaging applications. SPAD and CMOS platforms, meanwhile, offer scalability, portability, and design flexibility for high-density cortical mapping and relative blood flow monitoring. A hybrid approach could balance depth sensitivity, spatial coverage, and cost-effectiveness, supporting application-specific system designs optimized for both clinical and research settings.

## Funding and Acknowledgment

The authors acknowledge the funding support from NIH R01 (NIBIB Brain Initiative (7R01EB031759-03). We also thank Nick Bertone (Picoquant Inc.) for providing the demo unit of HydraHarp400 for the time domain acquisition system. M.W., P.M. and A.U. acknowledge partial support by the Swiss National Science Foundation (grants 20QT21_187716 Qu3D “Quantum 3D Imaging at high speed and high resolution” and 200021_166289), and in part by Meta Platforms Inc.

## Data availability

Data underlying the results presented in this paper are not publicly available at this time but may be obtained from the authors upon reasonable request.

